# Prognostic factors for severity and mortality in patients infected with COVID-19: A living systematic review protocol

**DOI:** 10.1101/2020.04.08.20056598

**Authors:** Ariel Izcovich, Martín Ragusa, Verónica Sanguine, Fernando Tortosa, Federico Espinosa, Camila Agnoletti, María Andrea Lavena Marzio, Agustina Ceirano, Agustín Bengolea, Ezequiel Saavedra, Hugo N. Catalano, Gabriel Rada, COVID-19 L·OVE Working Group

**Affiliations:** Servicio de clinica medica del Hospital Aleman, Buenos Aires, Argentina; Servicio de clinica medica del Hospital Fernandez, Buenos Aires, Argentina; Dirección Nacional de Calidad en Servicios de Salud y Regulación Sanitaria- Ministerio de Salud de la Nación, Argentina; Departamento Médico. Hospital “Ramón Carrillo”, San Carlos de Bariloche, Rio Negro, Argentina; Coordinación de evaluación de tecnologías. Ministerio de Salud de Rio Negro, Rio Negro, Argentina; Fundación Epistemonikos, Santiago, Chile; UC Evidence Center, Cochrane Chile Associated Center, Pontificia Universidad Católica de Chile, Santiago, Chile; Internal Medicine Department, Faculty of Medicine, Pontificia Universidad Católica de Chile, Santiago, Chile

**Keywords:** COVID-19, severe acute respiratory syndrome coronavirus 2, Coronavirus Infections, Systematic review, prognosis

## Abstract

**Objective:** The objective of our systematic review is to identify prognostic factors that can potentially be used in decision-making related to the care of patients infected with COVID-19.

**Design:** This is the protocol of a living systematic review.

**Data sources:** We will conduct searches in PubMed/Medline, Embase, Cochrane Central Register of Controlled Trials (CENTRAL), grey literature and in a centralised repository in L·OVE (Living OVerview of Evidence). L·OVE is a platform that maps PICO questions to evidence from Epistemonikos database. In response to the COVID-19 emergency, L·OVE was adapted to expand the range of evidence covered and customised to group all COVID-19 evidence in one place. The search will continue until the day before submission to a journal.

**Eligibility criteria for selecting studies and methods:** We will follow a common protocol for multiple parallel systematic reviews, already published and submitted to PROSPERO (awaiting ID allocation).

We will include studies that assess patients with confirmed or suspected SARS-CoV-2 infection and inform the relation between potential prognostic factors and death or disease severity. Two groups of two reviewers will independently screen studies for eligibility, extract data and assess the risk of bias. We will perform meta-analyses and use GRADE to assess the certainty of evidence for each prognostic factor and outcome.

A living, web-based version of this review will be openly available during the COVID-19 pandemic. We will resubmit it if the conclusions change or there are substantial updates.

**Ethics and dissemination:** No ethics approval is considered necessary. The results of this review will be widely disseminated via peer-reviewed publications, social networks and traditional media.

**PROSPERO Registration:** Submitted to PROSPERO (awaiting ID allocation).

## INTRODUCTION

COVID-19 is an infection caused by the SARS-CoV-2 coronavirus [1]. It was first identified in Wuhan, China, on December 31, 2019 [2]; three months later, almost half a million cases of contagion had been identified across 197 countries [3]. On March 11, 2020, WHO characterised the COVID-19 outbreak as a pandemic [1].

While the majority of cases result in mild symptoms, some might progress to pneumonia, acute respiratory distress syndrome and death [4],[5],[6]. The case fatality rate reported across countries, settings and age groups is highly variable, but it would range from about 0.5% to 10% [7]. In hospitalized patients it has been reported to be higher than 10% in some centres [8].

Prognostic factors (standalone or combined in risk assessment models) can be employed in patients with SARS-CoV-2 infection to stratify the different subsets of patients and their risk of developing severe disease or dying. This stratification may then support optimised management and resource utilization strategies to provide care to these patients.

Although multiple prognostic factors have been proposed and some are accepted as “established” by the scientific community (i.e age), the predictive capacity of most of these potential prognostic factors has not been properly evaluated and remains uncertain [9].

Using innovative and agile processes, taking advantage of technological tools, and resorting to the collective effort of several research groups, this living systematic review aims to provide a timely, rigorous and continuously updated summary of the evidence available on prognostic factors that can potentially be used in decision-making related to the care of patients infected with COVID-19.

## METHODS

### Protocol and registration

This manuscript complies with the ‘Preferred Reporting Items for Systematic reviews and Meta-Analyses’ (PRISMA) guidelines for reporting systematic reviews and meta-analyses [10]. Additionally we will follow the CHARM-PF checklist to determine which items to extract from primary studies of prognostic factors [11].

A protocol stating the shared objectives and methodology of multiple evidence syntheses (systematic reviews and overviews of systematic reviews) to be conducted in parallel for different questions relevant to COVID-19 was registered in PROSPERO (submitted, awaiting PROSPERO ID allocation) and published [12]. The protocol was adapted to the specificities of the question assessed in this review.

### Search strategies

#### Electronic searches

Our literature search was devised by the team maintaining the L·OVE platform (https://app.iloveevidence.com), using the following approach:

1. Identification of terms relevant to the population and intervention components of the search strategy, using Word2vec technology [13] to the corpus of documents available in Epistemonikos Database.
2. Discussion of terms with content and methods experts to identify relevant, irrelevant and missing terms.
3. Creation of a sensitive boolean strategy encompassing all of the relevant terms
4. Iterative analysis of articles missed by the boolean strategy, and refinement of the strategy accordingly.

Our main search source was Epistemonikos database (https://www.epistemonikos.org), a comprehensive database of systematic reviews and other types of evidence [14]. We supplemented it with articles from multiple sources relevant to COVID-19 (without any study design, publication status or language restriction) [15].

In sum, Epistemonikos Database acts as a central repository. Only articles fulfilling Epistemonikos criteria are visible by users. The remaining articles are only accessible for members of COVID-19 L·OVE Working Group.

Additional searches will be conducted using highly sensitive searches in PubMed/MEDLINE, the Cochrane Central Register of Controlled Trials (CENTRAL) and Embase, without any language or publication status restriction. The searches will cover from the inception date of each database until the day before submission.

The following strategy will be used to search in Epistemonikos Database. We will adapt it to the syntax of other databases.

(coronavir* OR coronovirus* OR “corona virus” OR “virus corona” OR “corono virus” OR “virus corono” OR hcov* OR “covid-19” OR covid19* OR “covid 19” OR “2019-nCoV” OR cv19* OR “cv-19” OR “cv 19” OR “n-cov” OR ncov* OR “sars-cov-2” OR “sars-cov2” OR (wuhan* AND (virus OR viruses OR viral)) OR (covid* AND (virus OR viruses OR viral)) OR “sars-cov” OR “sars cov” OR “sars-coronavirus” OR “severe acute respiratory syndrome” OR “mers-cov” OR “mers cov” OR “middle east respiratory syndrome” OR “middle-east respiratory syndrome” OR “covid-19-related” OR “SARS-CoV-2-related” OR “SARS-CoV2-related” OR “2019-nCoV-related” OR “cv-19-related” OR “n-cov-related”) AND (incidence* OR “follow-up” OR “follow up” OR prognos* OR predict* OR course*)

#### Other sources

In order to identify articles that might have been missed in the electronic searches, we will do the following:

1. Screen the reference lists of other systematic reviews, and evaluate in full text all the articles they include.
2. Scan the reference lists of selected guidelines, narrative reviews and other documents.
3. Conduct cross-citation search in Google Scholar and Microsoft Academic, using each included study as the index reference.
4. Review the reference list of each included study.

### Eligibility criteria

#### Types of studies

We will include prognostic factors and risk assessment model studies that are based on the typologies of prognosis proposed by Iorio and colleagues [16], based on the PROGnosis RESearch Strategy (PROGRESS) Group framework [17].

We will exclude studies evaluating effects on animal models or in vitro conditions.

#### Types of participants

We will include studies that evaluate patients with confirmed infection with SARS-CoV-2 independently of the healthcare setting (i.e ambulatory or inpatients) and the severity of infection.

We will exclude studies if the population did not reflect the general population of interest, such as studies that included patients with other viral infections (e.g. SARS-CoV or MERS-CoV).

#### Type of interventions

We will investigate all prognostic factors reported in individual studies and compare patients exposed with patients unexposed to each one of those factors. We will explore the prognostic information provided by every prognostic factor independently of ‘established’ prognostic factors (age, comorbidities and disease severity at the start of follow up).

#### Type of outcomes

We will not use outcomes as an exclusion criteria during the selection process. Any article meeting all the criteria except for the outcome criterion will be preliminarily included and evaluated in full text.

We will include studies that assess mortality or severe COVID-19 infection. We will accept the author’s definitions of severe COVID-19 infection. Additionally, we will consider all the outcomes that can be assumed to reflect COVID-19 severity (i.e ICU requirement).

#### Timing and setting

We will consider studies that provide information on prognostic factors measured at any time after the COVID-19 infection has been confirmed, independently of the healthcare setting (outpatient, general ward and ICU).

### Selection of studies

The results of the literature search in Epistemonikos database will be automatically incorporated into the [L·OVE platform](https://test.iloveevidence.com/loves/5e6fdb9669c00e4ac072701d) (automated retrieval), where they will be de-duplicated by an algorithm comparing unique identifiers (database ID, DOI, trial registry ID), and citation details (i.e. author names, journal, year of publication, volume, number, pages, article title and article abstract).

The additional searches will be uploaded to the screening software Collaboratron™ [18].

In both L·OVE platform and Collaboratron™, two researchers will independently screen the titles and abstracts yielded by the search against the inclusion criteria. We will obtain the full reports for all titles that appear to meet the inclusion criteria or require further analysis to decide on their inclusion.

We will record the reasons for excluding trials in any stage of the search and outline the study selection process in a PRISMA flow diagram adapted for the purpose of this project.

Two reviewers (AI and MR) will screen independently and in duplicate the titles and abstracts of all the retrieved citations; discrepancies will be resolved by consensus. They will then retrieve the full texts of all citations judged as potentially eligible. Two teams of two reviewers (AI-VS and MR-FT) will screen the full texts independently and in duplicate and compare results; disagreements will be resolved by consensus. Reviewers will use standardized screening forms and conduct calibration exercises before the screening process.

### Extraction and management of data

Two groups of two reviewers will abstract data independently and in duplicate, from all eligible studies, using standardized forms. Reviewers will compare and discuss results in case of any disagreement. We will conduct calibration exercises and pilot all forms, prior to the start of the data abstraction process.

For all identified studies - risk assessment model studies and prognostic factor studies - the reviewers will abstract data about the following characteristics:

- Study context (e.g. country, year of publication)
- Study design (e.g. cohort or case control; duration of follow up)
- Population and their demographics (e.g. sample size, age, number of centers)
- Outcomes definition, method of measurement and time of occurrence and missing information
- Prognostic factors, definitions, and measurement methods (including thresholds used for continuous predictors)
- Measures of association (e.g. odds ratio (OR)) or crude estimates (calculated from the number of patient exposed and non-exposed with and without the event)

### Risk of bias assessment

Two reviewers independently and in duplicate will assess risk of bias of the individual included studies. Discrepancies will be resolved by consensus. We will assess the risk of bias in the included studies using the Quality in Prognosis Studies tool (QUIPS) for prognostic factor studies [19]. In order to assess adequacy of the multivariate model used to adjust effect estimates we will consider as appropriate the models that include all of the following:

- Age
- One comorbidity (i.e diabetes)
- One parameter of disease severity (i.e disphnea)

### Measures of treatment effect

For every individual prognostic factor, we will present the measure of association with the corresponding outcome. We will present the effect estimate as odds ratios (ORs) and their corresponding 95% confidence intervals (CI). In studies that report the measure of association as a hazard ratio (HR) or risk ratio (RR), we will convert them to ORs using the baseline risk (death rate or incidence of severe COVID-19 infection out of the total sample) reported in the studies [20], [21]. We will use the crude effect estimates when the adjusted estimates are not provided.

### Strategy for data synthesis

We will present the results of the included studies including the individual prognostic factors in both tabular and narrative formats. We will standardize the units of measurement for each prognostic factor, unifying the direction of the predictors and adjusting the weights of the studies, and calculate crude effect estimates when not provided [22]. When possible, we will meta-analyze all prognostic factors that are both associated with the outcomes mortality and severe COVID-19 infection and reported by more than one study. We will use the generic inverse variance-based method to produce an overall measure of association. We will explore consistency of the associations between our meta-analyzed results and studies reporting the same predictors that could not be pooled. We will use random-effect models provided by the metafor package for R software [23].

### Subgroup and sensitivity analysis

To explore the robustness of the estimates of effects of every prognostic factor and for every outcome we will perform the following additional analysis:

Subgroup analysis: We will perform subgroup analysis according to the definition of severe COVID-19 infection (i.e respiratory failure vs respiratory distress syndrome vs ICU requirement). In case we identify significant differences between subgroups (test for interaction <0.05) we will report the results of individual subgroups separately.

Sensitivity analysis: We will perform sensitivity analysis excluding high risk of bias studies and studies that did not report adjusted estimates. In cases where the effect estimates provided by the primary analysis and the sensitivity analysis significantly differ, we will either present the low risk of bias – adjusted estimates or present the primary analysis estimates but rate down certainty of the evidence because of risk of bias.

If necessary, we will perform other subgroup and sensitivity analyses accounting for alternative ways of prognostic factors/outcome measurement or presentation.

### Assessment of certainty of evidence

For every individual prognostic factor, we will present the measure of association with the corresponding outcome. We will perform an assessment of the certainty of evidence for each of the prognostic factors, per outcome, based on the GRADE approach [24]. The approach considers the following domains: risk of bias, indirectness, inconsistency, imprecision, and publication bias. We will develop summary of findings tables and rate the overall certainty of evidence as high, moderate, low or very low depending on the grading of the individual domains [24].

### Living evidence synthesis

An artificial intelligence algorithm deployed in the Coronavirus/COVID-19 topic of the L·OVE platform (https://app.iloveevidence.com/loves/5e6fdb9669c00e4ac072701d) will provide instant notification of articles with a high likelihood to be eligible. The authors will review these and will decide upon inclusion, and will update the [living web version](https://app.iloveevidence.com/living-reviews/5e7e748c53e53e78053563fd) of the review accordingly. We will consider resubmission to a journal if there is a change in the direction of the effect on the critical outcomes or a substantial modification to the certainty of the evidence.

This review is part of a larger project set up to produce multiple parallel systematic reviews relevant to COVID-19 [12].

## Data Availability

The data that will support the findings of this study will be openly available.

## NOTES

## Acknowledgements

The members of the COVID-19 L·OVE Working Group and [Epistemonikos Foundation](https://www.epistemonikos.cl/) have made it possible to build the systems and compile the information needed by this project. Epistemonikos is a collaborative effort, based on the ongoing volunteer work of over a thousand contributors since 2012.

## Roles and contributions

GR conceived the common protocol for all the reviews being conducted by the COVID-19 L·OVE Working Group. AI and GR drafted the manuscript, and all other authors contributed to it. The corresponding author is the guarantor and declares that all authors meet authorship criteria and that no other authors meeting the criteria have been omitted.

The COVID-19 L·OVE Working Group was created by Epistemonikos and a number of expert teams in order to provide decision makers with the best evidence related to COVID-19. Up-to-date information about the group and its member organisations is available here: epistemonikos.cl/working-group

## PROSPERO registration

This protocol has been submitted (awaiting PROSPERO ID allocation).

## Ethics

As researchers will not access information that could lead to the identification of an individual participant, obtaining ethical approval was waived.

## Data sharing

All data related to the project will be available. Epistemonikos Foundation will grant access to data.

